# Magnetic resonance imaging and the evaluation of vestibular schwannomas: a systematic review

**DOI:** 10.1101/2025.06.06.25329105

**Authors:** Keng Siang Lee, Navodini Wijetilake, Steve Connor, Tom Vercauteren, Jonathan Shapey

## Abstract

**Introduction:** The assessment of vestibular schwannoma (VS) requires a standardized measurement approach as growth is a key element in defining treatment strategy for VS. Volumetric measurements offer higher sensitivity and precision, but existing methods of segmentation, are labour-intensive, lack standardisation and are prone to variability and subjectivity. A new core set of measurement indicators reported consistently, will support clinical decision-making and facilitate evidence synthesis. This systematic review aimed to identify indicators used in 1) magnetic resonance imaging (MRI) acquisition and 2) measurement or 3) growth of VS. This work is expected to inform a Delphi consensus.

**Methods:** Systematic searches of Medline, Embase and Cochrane Central were undertaken on 4^th^ October 2024. Studies that assessed the evaluation of VS with MRI, between 2014 and 2024 were included.

**Results:** The final dataset consisted of 102 studies and 19001 patients. Eighty-six (84.3%) studies employed post contrast T1 as the MRI acquisition of choice for evaluating VS. Nine (8.8%) studies additionally employed heavily weighted T2 sequences such as constructive interference in steady state (CISS) and FIESTA-C. Only 45 (44.1%) studies reported the slice thickness with the majority 38 (84.4%) choosing <3mm in thickness. Fifty-eight (56.8%) studies measured volume whilst 49 (48.0%) measured the largest linear dimension; 14 (13.7%) studies used both measurements. Four studies employed semi-automated or automated segmentation processes to measure the volumes of VS. Of 68 studies investigating growth, 54 (79.4%) provided a threshold. Significant variation in volumetric growth was observed but the threshold for significant percentage change reported by most studies was 20% (n = 18).

**Conclusion:** Substantial variation in MRI acquisition, and methods for evaluating measurement and growth of VS, exists across the literature. This lack of standardization is likely attributed to resource constraints and the fact that currently available volumetric segmentation methods are very labour-intensive. Following the identification of the indicators employed in the literature, this study aims to develop a Delphi consensus for the standardized measurement of VS and uptake in employing a data-driven artificial intelligence-based measuring tools.

## Introduction

Vestibular schwannoma (VS) is a benign brain tumour originating from peripheral myelinating Schwann cells within the vestibular division of the vestibulocochlear nerve. The advent availability and improved quality of magnetic resonance imaging (MRI) has resulted in higher prevalence of small asymptomatic VS now being diagnosed and patients typically require an extended period of surveillance.^1^

The assessment of VS growth, which is a key element in defining treatment strategy for VS, requires a standardized measurement approach.^2^ The 2003 International Consensus Meeting on Systems for Reporting Results in VS recommends distinguishing intrameatal and extrameatal portions and defining size of the VS by its maximal linear diameter.^3^ However, linear measurements may give the impression of intermittent growth, which complicates clinical decision-making. On the contrary, volumetric measurement is a more sensitive and precise method of calculating the true size of the VS and is superior at detecting subtle growth.^4,5^ Implementing routine volumetric measurements would enable clinicians to offer earlier appropriate intervention. However, existing methods, including manual or semi-automated tumour segmentation, are labour-intensive, lack standardisation and are prone to subjectivity and hence variability.^6^ Additionally, there is a lack of readily available dedicated software for routine clinical practice.^7^ To manage this implementation gap, we have previously developed a fully automated artificial intelligence (AI) framework to segment VS using MRI, achieving state-of-the-art capability to fully automate the detection and segmentation of VS, and can also delineate the tumour’s intra and extra-meatal components.^7–9^

However, there is a need to develop a consistent approach for reporting the indicators used to inform clinical decision making for patients with VS. A revised core set of measurement indicators reported consistently, will support clinical decision-making and facilitate evidence synthesis. To inform a Delphi consensus, we undertook a systematic review to identify potential indicators used in current clinical practice for the 1) MRI acquisition or evaluation of 2) measurement or 3) growth of VS.

## Methods

This review was conducted according to the Preferred Reporting Items for Systematic Reviews and Meta-Analyses (PRISMA) guidelines.^10^ The protocol was registered on the PROSPERO international prospective register of systematic reviews (registration number CRD42024604452).^11–13^

### Search strategy

Searches of three electronic databases were undertaken including Ovid Medline, Ovid Embase, and Cochrane Central Register of Controlled Trials (CENTRAL). Searches were performed in each database from its inception until 4^th^ October 2024. The concepts of “MRI”, “acquisition”, “measurement”, “growth” and “VS”, were used in addition to synonyms and related terms. The full search strategy used for the databases is presented in Supplementary Table 1.

### Study selection

All titles and abstracts were screened against the pre-defined eligibility criteria developed independently by two reviewers (KSL and NW). A full list of inclusion and exclusion criteria can be found in Supplementary Table 2. Disagreements were resolved by discussion, and where agreement could not be reached, the senior reviewer assisted with decision making (JS).

In the event of multiple publications analyzing the same cohort, the publication that reported the greatest number of relevant data was used for evaluation. This was to avoid multiple counting which overstates sample size, leading to an artificially exaggerated precision in the summary estimate.^11–13^

### Data extraction

A pro forma was developed and piloted to extract data on the following variables to ensure standardization and consistency in this process: 1) study details, 2) MRI acquisition, and 3) methods for evaluating measurement or growth of VS.

### Data analysis and reporting

A narrative synthesis of data, with descriptive analyses where appropriate, was undertaken. A risk of bias assessment of studies or meta-analysis was not performed as the aim of this review was only to identify indicators to inform a Delphi consensus, and not to appraise the validity of their findings.^12,13^

## Results

### Study selection

Following screening of 2115 unique articles, 102 studies including 19001 patients, reporting data on MRI acquisition and the evaluation of measurement or growth of VS, were included in the final dataset (Figure 1). Eighty-three studies were excluded because they had reported data from the same cohort of patients across overlapping time periods. These were mostly from institutions such as the Mayo Clinic Rochester,^14–21^ and the University of Pittsburgh Medical Center.^22–25^

**Figure 1.**
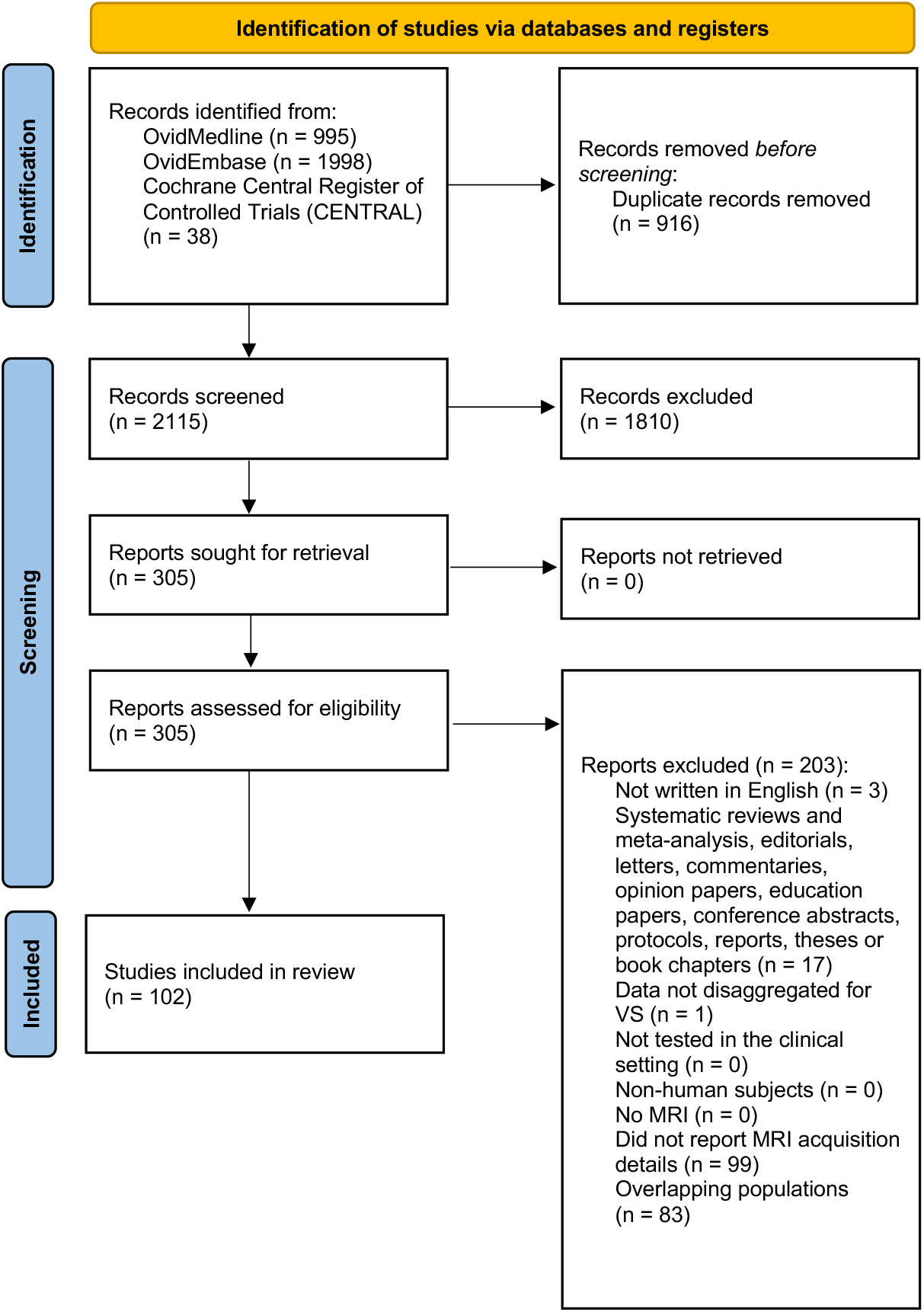
PRISMA flow diagram for studies included and excluded from the systematic review

**Figure 2.**
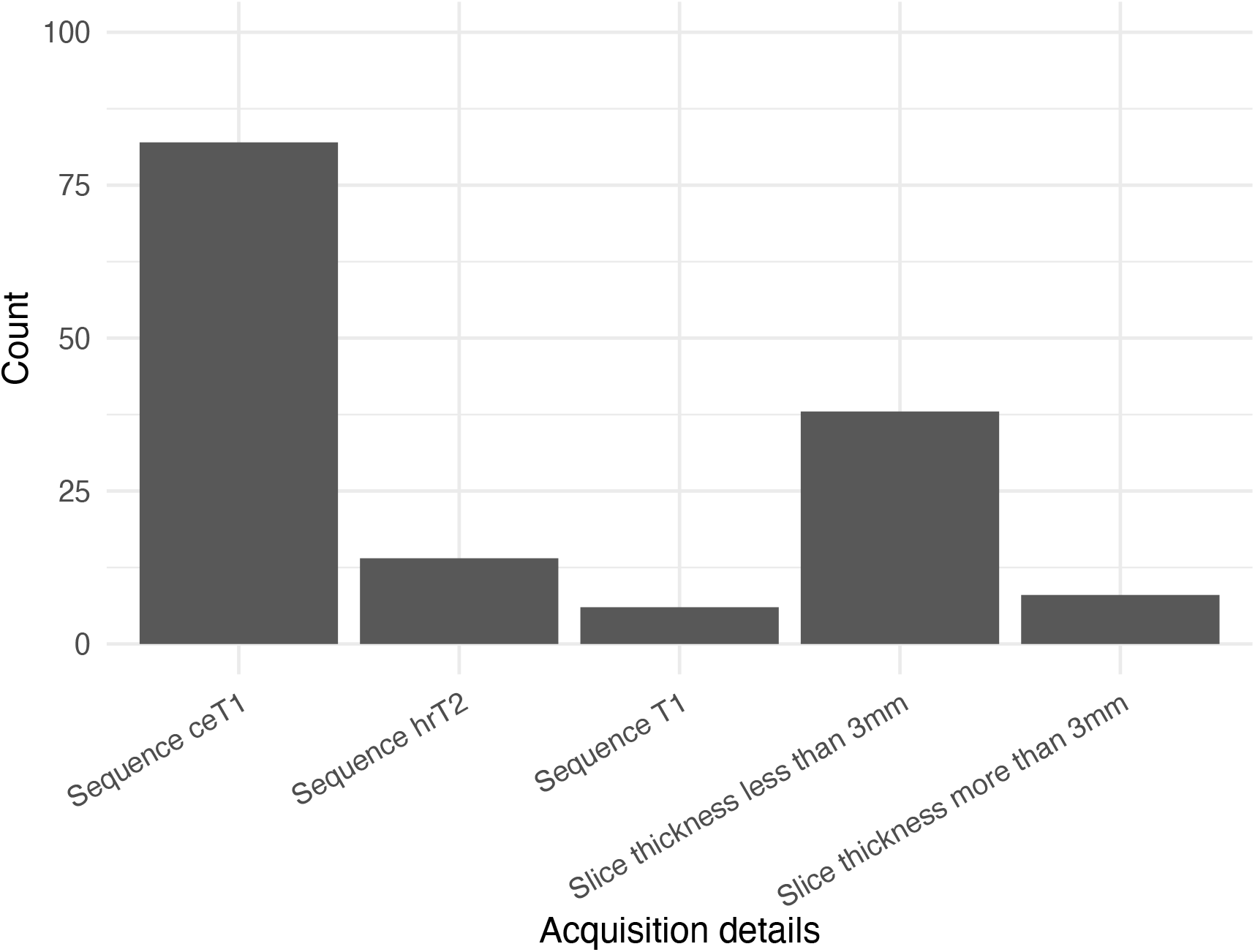
The most reported sequences amongst the studies. ceT1 = contrast enhanced T1. hrT2 = high resolution T2

### Characteristics of included studies

Due to the volume of included full texts, a complete list of all characteristics synthesised from the 102 primary studies together with their corresponding Digital Object Identifiers (DOIs) are shown in Supplementary Table 3.

Data were collected across 25 countries: 26 from the United States; nine from the United Kingdom; seven each from France, Germany, Japan and South Korea; five each from China, Italy and Taiwan; four from the Netherlands; three from Australia; two each from Belgium, India and Spain; and one each from Austria, Brazil, Canada, Denmark, Dominican Republic, Hong Kong, Norway, Poland, Singapore, Turkey, and Switzerland.

### Indicators used for the acquisition of MRI

The majority 86 (84.3%) of studies employed contrast-enhanced T1-weighted (ceT1) as the MRI acquisition of choice for evaluating VS. Nine (8.8%) studies additionally employed heavily weighted T2 sequences such as CISS (constructive interference in steady state) and FIESTA-C (fast imaging employing steady-state acquisition). Eight (7.8%) studies did not specify the MRI sequence but mentioned only the use of contrast enhancement. Only 45 (44.1%) studies reported the slice thickness with the majority of these (38 studies, 84.4%) choosing <3mm in thickness.

### Indicators used for the measurement of the VS

Fifty-eight (56.8%) studies measured tumour volume (TV) by estimation from linear measurements whilst 49 (48.0%) measured the maximal linear dimension (MLD). Fourteen (13.7%) studies used both measurements.

Four studies employed semi-automated to automated segmentation processes to measure the volumes of VS.

### Indicators used for evaluation of growth of the VS

Sixty-eight (66.7%) studies investigated VS growth and reported the indicators used for its evaluation. Of these 68 studies, 54 (79.4%) provided a threshold for defining growth. Significant variation in defining growth based on linear dimensions exist. In most studies (n = 21), the MLD threshold used to define definitive growth was 2mm over the imaging interval. Less frequently reported threshold was a percentage change of 10% (n = 2).

Significant variation in volumetric growth was observed but the threshold for significant percentage change reported by most studies was 20% (n = 18). Less frequently reported threshold was a percentage change of 10% (n = 11) or 25% (n = 2), or absolute change of >100mm^3^/year (n = 1) or >0.05cm^3^ between two consecutive MRIs (n = 1).

Absolute volumetric change was rarely employed (n=2) but ranged 100-133mm^3^ per time interval. Other studies cited Response Assessment in Neuro-Oncology (RANO) (n=1),^26^ Response Evaluation Criteria in Solid Tumors (RECIST) (n=3),^27^ and Response Evaluation in Neurofibromatosis and Schwannomatosis International Collaboration (REiNS) (n=1) criteria,^28^ in determining their threshold for volumetric growth.

## Discussion

### Summary of findings

This systematic review demonstrated substantial variation in the MRI acquisition, and methods for evaluating measurement and growth of VS, across in the literature.

### Implications for research and practice

Heterogeneous methods used to inform the management of VS limits the extent to which evidence synthesis using data from the literature can provide valid information about the effectiveness of treatments for VS.^29–31^

Whilst it is accepted that MRI is the standard of care for the imaging of VS, the choice of MRI sequences will depend on whether they are being used for detection, characterisation or follow up and there is international variability in practice. Fluid sensitive 3D gradient echo (GRE) balanced steady state free precession (e.g. CISS, FIESTA-C) or contemporary T2w 3D fast spin echo (FSE) (e.g. SPACE,CUBE) techniques are widely used for the detection of vestibular schwannomas in patients with audio-vestibular symptoms. In many centres, T1w 3D sequences pre and post gadolinium are only performed if there is an equivocal finding or in clinical scenarios where alternative pathologies are more likely (e.g. immunosuppression or malignancy). As observed from our study, the majority of studies (84.3%) employed ceT1 as the MRI acquisition of choice for evaluating VS. However, some guidelines also recommend the routine use of T1w 3D sequences for detection,^32^ and there are potential benefits in the detection of lesions < 2 mm in size.^33^ Following detection of a cerebello-pontine angle cistern or internal auditory canal lesion, pre and post gadolinium T1w 3D sequences would routinely be used for characterisation in order to distinguish from other tumour mimics. Post gadolinium imaging may also help delineate cystic changes and distinguish tumour from obstructed fluid in the fundus of the internal auditory canal.

The most cost-effective monitoring of the untreated VS is performed with T2w 3D sequences alone,^34^ however pre and post gadolinium T1w 3D sequences are usually performed following therapeutic interventions to better delineate residual and recurrent tumor from post-operative scar tissue and post-treatment change. Fat saturated T1w imaging may be a useful adjunct in the setting of a post-operative fat graft. The delineation of tumour contour for serial volumetric analysis is generally superior with post gadolinium T1w 3D sequences due to the greater contrast to noise ratio. The precise contrast characteristics of the T2w 3D sequences is variable and it may potentially result in suboptimal delineation of tumour from the adjacent bone or brain.^35^ It should be noted that the slice thickness influences the volumetric evaluation of tumour size, and whilst older 2D FSE sequences were generally limited to 2-3 mm slice thickness, thinner sections are better achieved with 3D T1w sequences. Since the mean IAC diameter is 5-6 mm, and the volumetric analysis based on fewer than five slices yields unacceptably larger errors,^36^ a maximum 1 mm slice thickness, is recommended since it can be better applied to the evaluation of intracanalicular tumours.^37^

The 2003 International Consensus Meeting on Systems for Reporting Results in VS has attempted to standardize VS reporting to facilitate evidence syntheses to provide more valid information about treatment effects.^3^ Due to the lack of available resources two decades ago, the meeting had recommended using maximal linear measurements to guide clinical management. Since then, studies have shown that volumetric measurement is a more sensitive and precise method of calculating the true size of the VS and is superior at detecting subtle growth.^4,5^ However, existing methods, including manual or semi-automated tumour segmentation, are labour-intensive, lack standardization and are prone to variability. These factors have hindered the adoption of incorporating volumetric measurements into routine clinical practice.^7,38,39^ In this systematic review, 56.8% of studies measured TV by estimating TV from linear measurements.

In recent years, various research groups have sought to address the practical limitations of the previously developed consensus-based criteria by developing automated segmentations methods that would facilitate the routine clinical use of volumetric measurements in the management of patients with VS.^39^ We previously developed the first fully automated AI pipeline to segment VS from MRI images.^7–9^ The method was trained and validated using ceT1 and high-resolution T2-weighted (hrT2) images from a dataset of 242 consecutive patients treated with Gamma knife SRS. Excellent accuracy scores were achieved (mean dice scores >93%), comparable to repeated measurements performed by clinicians.^8^ Since then, to address the domain shift, further robust well-generalized models have been developed and validated to track tumour growth, thereby augmenting clinical decision-making.^38,40–48^ AI-driven clinical support tools have the potential to improve patient outcomes and experience by the standardization and personalization of VS treatment. AI-driven technology is expected to facilitate quicker and more objective treatment decisions to be made and may safely reduce the number of scan and clinic appointments.^38^ However, the acceptance of AI technology in clinical decision-making and patient care remains a barrier to clinical translation.^49–53^ Continued engagement with clinicians and patients is therefore required to ensure that the benefits of AI-driven technologies are fully realized.

### Future directions

Through a pragmatic multiphase approach, we plan to conduct a Delphi consensus in collaboration with the British Skull Base Society (BSBS) to describe how AI and advanced medical technology may be used to support the management of patients with VS. Informed by this systematic review, we aim to establish a consensus for the optimal MRI acquisition(s) for VS and will explore which segmentation-based measurements – including volumetric and extracted linear measurements – should be used to determine tumour size and growth. The Delphi consensus will also explore the requirements of clinicians towards the integration of AI into patient management including potential user interfaces, its integration into the planning of radiation treatment and the technology’s explainability and trustworthiness. We will engage with a steering group representing relevant multidisciplinary stakeholders to develop the specific questions for the Delphi study and we will encourage active participation in the Delphi process by a wide group of relevant clinical stakeholders in the hope of ensuring future clinical uptake of the resulting consensus.

## Limitation

This systematic review employed a pre-specified, registered protocol.^13^ The literature search was comprehensive, identifying relevant studies from three databases, and the reporting of this study follows PRISMA guidelines (Supplementary Table 4 for PRISMA checklist).^11^ MRI technology has advanced considerably over the last decade and although this reflects modern practice, could have confounded the results. Limitations of this systematic review are that we were only able to include publications written in English, due to resource constraints. Therefore, it is possible that some relevant studies were omitted. However, international publications were included to reduce the risk of selection bias.^11^

## Conclusion

This systematic review demonstrates substantial variation in the 1) MRI acquisition, and methods for evaluating 2) measurement and 3) growth of VS, across in the literature. This heterogeneity may potentially hinder effective evidence synthesis and limits the validity of evidence available for the identification of effective treatments for VS. This lack of standardization is likely attributed to resource constraints and currently available labour-intensive methods for automated tumour segmentation. With new the advent use of technology, this study aims to reach a consensus to standardize the workflow to managing VS. This will result in consistent approach employing AI and recording of data about VS across studies and will allow the accumulation and comparison of evidence to identify the most effective treatments for patients with VS. With identification of the indicators employed in the literature, this systematic review will inform a Delphi consensus to reach a standardized approach for the evaluation of VS.

## Data Availability

Upon request from corresponding author

## Declarations

### Ethics and Consent to Participate declarations

Not applicable

### Author contributions

Authors’ contributions (CRediT) All authors listed have made substantial, direct, and intellectual contribution to the work and approved it for publications.

KSL: Conceptualization, Methodology, Formal analysis and investigation, Writing - original draft preparation, Writing - review and editing, Visualization, Funding acquisition.

NW: Data curation

SC: Writing - review and editing

TC: Writing - review and editing, Supervision.

JS: Conceptualization, Writing - review and editing, Supervision, Funding acquisition.

## Abbreviation list

ceT1: contrast enhanced T1
CISS: constructive interference in steady state
FIESTA: fast imaging employing steady-state acquisition
hrT2: high resolution T2
MLD: maximal linear diameter
VS: vestibular schwannomas

