## Supplementary tables 1, 2 and 3 for "Magnetic resonance imaging and the evaluation of vestibular schwannomas: a systematic review"

Supplementary Table 1. Search strategy used for the three electronic databases 4 October 2024

| **EMBASE search** | **1998 articles** |
|---|---|
| No. | Search term |
| **Vestibular schwannoma concept** | |
| 1 | exp acoustic neuroma/ |
| 2 | (vestibular schwannoma* or acoustic neuroma*).tw. |
| 3 | 1 or 2 |
| **MRI concept** | |
| 4 | exp nuclear magnetic resonance imaging/ |
| 5 | (magnetic resonance imaging or MRI).tw. |
| 6 | 4 or 5 |
| **Outcome concept** | |
| 7 | (linear or rate or progression or growth or analysis or volume* or 3D* or size or segment* or diameter* or acquisition or sequence* or measure*).tw. |
| **Combined concepts** | |
| 8 | 3 and 6 and 7 |
| 9 | limit 8 to yr="2014 -Current" |
| **OVID Medline search** | **995 articles** |
| No. | Search term |

| | |
|---|---|
| **Vestibular schwannoma concept** | |
| 1 | exp Neuroma, Acoustic/ / |
| 2 | (vestibular schwannoma* or acoustic neuroma*).tw. |
| 3 | 1 or 2 |
| **MRI concept** | |
| 4 | exp magnetic resonance imaging/ |
| 5 | (magnetic resonance imaging or MRI).tw. |
| 6 | 4 or 5 |
| **Outcome concept** | |
| 7 | (linear or rate or progression or growth or analysis or volume* or 3D* or size or segment* or diameter* or acquisition or sequence* or measure*).tw. |
| **Combined concepts** | |
| 8 | 3 and 6 and 7 |
| 9 | limit 8 to yr="2014 -Current" |

| **Cochrane Central Register of Controlled Trials (CENTRAL)** | **38 articles** |
|---|---|
| No. | Search term |
| **Vestibular schwannoma concept** | |
| 1 | MeSH descriptor: [Neuroma, Acoustic] explode all trees |
| 2 | (vestibular schwannoma* or acoustic neuroma*):ti,ab,kw |
| 3 | #1 or #2 |

| | |
|---|---|
| **MRI concept** | |
| 4 | MeSH descriptor: [magnetic resonance imaging] explode all trees |
| 5 | (magnetic resonance imaging or MRI):ti,ab,kw |
| 6 | #4 or #5 |
| **Outcome concept** | |
| 7 | (linear or rate or progression or growth or analysis or volume* or 3D* or size or segment* or diameter* or acquisition or sequence* or measure*):ti,ab,kw |
| **Combined concepts** | |
| 8 | #3 and #6 and #7 |

Supplementary Table 2. Inclusion and exclusion criteria used to select studies for the review

| **Inclusion criteria** | **Exclusion criteria** |
|---|---|
| Primary MRI surveillance studies assessing/reporting the 1) acquisition OR evaluation of 2) measurement, OR 3) growth of the VS | • Not written in English<br>• Systematic reviews and meta-analyses, editorials, commentaries, opinion papers, letters, education papers, conference abstracts, protocols, reports, theses or book chapters<br>• Not exclusively about VS (either not about VS at all or about VS within a heterogenous CPA population)<br>• Not tested in the clinical setting (e.g. lab based rather than clinical practice)<br>• Non-human subjects (e.g. murine, porcine studies)<br>• No MRI<br>• Did not report indicators of interest (e.g. neither acquisition, or measurement or growth)<br>• Overlapping populations |

Supplementary Table 3. Characteristics of the 103 included studies and the indicators used for the acquisition of MRI, measurement and growth of VS.

| First author | Year | DOI | Country | Data collection period | Study design | No. of patients | Primary aim of study | Methods of MRI acquisition | Indicators for measurement | Indicators for growth |
|---|---|---|---|---|---|---|---|---|---|---|
| Aktan SL | 2022 | 10.7759/cureus.22231 | USA | 1995-2019 | Case series | 372 | To evaluate the growth patterns of very small intracanalicular VS measuring ≤ 4 mm in MLD | ceT1 [x] | MLD | MLD ≥2mm |
| Albano L | 2023 | 10.3171/2022.7.JNS22812 | USA | 2015-2019 | Retrospective cohort study | 234 | To evaluate volumetric response of VS following SRS | ceT1 [x] | TV | TV ≥20%<br>18 months (short-term follow-up) and up to 5 years (long-term follow-up) |
| Ali NES | 2023 | 10.1177/00034894221119613 | USA | 2008-2016 | Case series | 21 | To evaluate growth rates of cystic VS | T1 and T2 [x] | TV<br>MLD | MLD ≥2mm/year<br>TV ≥20% |
| Arambula AM | 2023 | 10.1097/MAO.0000000000003814 | USA | 2010-2019 | Retrospective cohort study | 43 | To compare the completeness of resection of VS according to surgical approach | hrT2[CISS]<br>hrT2[c-FIESTA]<br><1mm slice thickness | TV | Not analysed |
| Balossier A | 2024 | 10.1093/neuonc/noae187 | France | 1992-2017 | Retrospective cohort study | 1607 | To evaluate the post-GKRS dynamics of evolution of VS volume, and characterize volumetric patterns. | ceT1<br>1-1.5mm slice thickness | TV | Not analysed |
| Bathla G | 2016 | 10.1097/MAO.0000000000001150 | USA | 2009-2011 | Retrospective cohort study | 41 | To determine if 2D and volumetric imaging parameters in VS correlate with hearing loss at presentation | ceT1<br>3mm slice thickness | TV<br>MLD | Not analysed |
| Bojrab DI 2nd | 2021 | 10.1097/MAO.0 | USA | 2007-2017 | Retrospective | 106 | To evaluate the relationship between fundal fluid and hearing outcomes | ceT1<br>1mm slice | MLD | Not analysed |

| | | | | | | | | | | |
|---|---|---|---|---|---|---|---|---|---|---|
| | | 0000 0000 0002 837 | | | cohort study | | after treatment of VS with GKRS | hrT2[CISS] hrT2[c-FIESTA] x | | |
| Borsetto D | 2022 | 10.3171/2022.1.JNS211544 | UK & Italy | 1990-2010 | Retrospective cohort study | 354 | To evaluate the long-term conditional probability of new VS growth for patients after 5 years of demonstrated nongrowth | ceT1, hrT2[CISS] T2 1mm slice thickness | MLD | MLD ≥2mm |
| Bossi Zanetti I | 2023 | 10.3390/jpm13050808 | Italy | 2004-2016 | Retrospective cohort study | 108 | To predict VS response to radiosurgery by applying ML algorithms on radiomic features extracted from pre-treatment MRI | ceT1 1mm slice thickness | TV | No threshold |
| Bourque JM | 2024 | 10.1097/MAO.0000000000004275 | Australia | 1998-2023 | Retrospective cohort study | 430 | To describe the natural history of growing VS and identify prognostic factors | ceT1 T2 0.5-1.6mm slice thickness | MLD | MLD ≥2mm |
| Bowden G | 2017 | 10.1093/neuros/nyx027 | USA | 2003-2012 | Retrospective cohort study | 219 | To correlate the radiographic appearance of VS before radiosurgery with delayed volumetric response | ceT1 <3mm slice thickness | TV | Regression – TV >20% reduction Growth or pseudoprogression – TV >10% increase |
| Bozhkov Y | 2022 | 10.1177/01945998211012674 | Germany | 2014-2017 | Retrospective cohort study | 138 | To evaluate postoperative hearing preservation in VS surgery | ceT1 x | MLD | Not analysed |
| Cazzador D | 2017 | 10.1080/21695717.2017.1385333 | Italy | 2012-2017 | Retrospective cohort study | 108 | To evaluate tumour growth and hearing decay in patients diagnosed with VS submitted to a wait and scan protocol of treatment | ceT1 x | MLD | Growth – MLD >2mm increase |
| Cesme DH | 2021 | 10.2174/1573405617 | Turkey | 2014-2019 | Retrospective cohort study | 31 | To determine the change in total tumor volume (TTV) in terms of radiological response in patients who | Post-contrast 3D T1-MPRAGE x | TV | Regression – TV 20% Progression – TV >20% increase |

| | | | | | | | | | | |
|---|---|---|---|---|---|---|---|---|---|---|
| | | 666210127160848 | | | | | had VS and were treated with radiosurgery and to investigate the relationship between the TTV, follow-up times and DTI parameters. | | | Stable – TV <20% change |
| Chang HC | 2023 | 10.1007/s11060-023-04280-z | Taiwan | 2003-2020 | Retrospective cohort study | 43 | To quantify the degree of brain stem deformity to predict long-term outcomes of patients with large VS following GKRS | ceT1 [x] | TV | No threshold |
| Chang J | 2019 | 10.1002/lary.27427 | USA | 2002-2014 | Retrospective cohort study | 62 | To determine if volumetric growth prior to GKRS predicts long-term tumor control | ceT1<br>0.8-7mm slice thickness | TV | Growth – TV >20% |
| Chew CH | 2022 | 10.21037/tro-22-1 | Taiwan | 2002-2020 | Retrospective cohort study | 50 | To evaluate the outcome of VS patients treated with SRS with a reduced marginal dose | ceT1 [x] | TV | Regression – MLD >2mm reduction or TV >10% reduction<br>Stable – MLD <2mm and <10% reduction |
| Choi Y | 2018 | 10.3857/roj.2018.00031 | South Korea | 2010-2016 | Retrospective cohort study | 42 | To explore the feasibility of maximum diameter as a response assessment method for VS after SRS or fractionated SRT | ceT1<br><3mm slice thickness | TV<br>MLD | Complete response as disappearance of VS.<br><br>As per RECIST<br>Partial response – MLD 30% reduction<br>Progressive disease – MLD 20% increase<br><br>Partial response – TV 67% reduction<br>Progressive disease – TV 73% increase |
| Cornelissen S | 2024 | 10.1007/s00234-024-03416-w | Netherlands | 2005-2020 | Retrospective cohort study | 100 | To determine the tumor volume dependency on the limits of agreement for volumetric measurements of VS by means of an inter-observer study | ceT1<br>0.8-2.0mm slice thickness | TV | Not analysed |
| Currie S | 2019 | 10.1259/bjr.20180833 | UK | Since 2017 | Retrospective cohort study | 50 | To examine whether the model of Getting It Right First Time (GIRFT) could be relevant to the | ceT1<br>3mm slice thickness<br>hrT2 | MLD | Not analysed |

| | | | | | | | | | | |
|---|---|---|---|---|---|---|---|---|---|---|
| | | | | | | | surveillance of non-operated VS | | | |
| D'Haese S | 2019 | 10.1080/00016489.2019.16352 68 | Belgium | 2003-2015 | Retrospective cohort study | 62 | To evaluate the natural course of VS and predictive factors of growth | ceT1, if unavailable hrT2[CISS] 0.5-3mm slice thickness | MLD | MLD ≥2mm/year |
| Daoudi H | 2024 | 10.3171/2023.7.JNS23138 | France | 2016-2019 | Retrospective cohort study | 512 | To evaluate the evolution of sporadic VS with radiological signs of VS regression and to identify prognostic factors for tumor shrinkage | ceT1and T2 [x] | TV | Regression – TV >20% reduction or MLD ≥2mm reduction |
| De Leo AN | 2024 | 10.1097/COC.0000000000001065 | USA | 1995-2019 | Retrospective cohort study | 139 | To evaluate outcomes report from an unselected series of patients with VS with radiographic brainstem compression treated with LINAC SRS | ceT1 1mm slice thickness | MLD | No threshold |
| Dhayalan D | 2023 | 10.1001/jama.2023.12222 | Norway | 2014-2022 | RCT | 120 | To evaluate upfront radiosurgery, or a careful follow-up by MRI in VS | ceT1 [x] | TV | No threshold |
| Ellenbogen JR | 2015 | 10.3109/02688697.2015.1036837 | UK | 2003-2009 | Retrospective cohort study | 50 | To examine tumour control, via volume changes, and the complications of LINAC-based SRS treatment of VS on medium-term follow-up. | ceT1 [x] | TV | No threshold |
| Fayad JN | 2014 | 10.1097/MAO.0000000000000285 | USA | 1988-1996 | Retrospective cohort study | 114 | To evaluate long-term prevalence of tumor growth and need for further treatment in patients with VS treated with conservative management | ceT1 [x] | MLD | Growth – MLD ≥2mm increase |

| | | | | | | | | | | |
|---|---|---|---|---|---|---|---|---|---|---|
| Fieux M | 2020 | 10.1080/00016489.2020.17176088 | France | 2010-2018 | Retrospective cohort study | 450 | To model residual VS over time to identify prognostic factors of postsurgical growth | ceT1 x | TV | Growth – TV >0.05 $cm^3$ increase |
| Fink KR | 2022 | 10.1016/j.acra.2020.09.022 | USA | 2019-2020 | Retrospective cohort study | 20 | To compare the precision and reproducibility of three different radiographic measurement techniques for assessing VS tumor size | Post-contrast 3D T1-MPRAGE 1mm slice thickness | MLD | Not analysed |
| Forgues M | 2018 | 10.1017/S0022215118001342 | USA | 2008-2016 | Retrospective cohort study | 26 | To assess the feasibility of non-contrast T2-weighted MRI as compared to T1-weighted post-contrast MRI for detecting VS growth | ceT1 hrT2 x | MLD | Not analysed |
| Fouard O | 2022 | 10.1016/j.ctro.2021.12.003 | Belgium | 2008-2015 | Retrospective cohort study | 63 | To determine a time interval during which a true VS progression can be distinguished from a pseudoprogression | ceT1, if unavailable T2 x | TV MLD | Change – TV 13% variation from baseline Pseudoprogression – TV 10% and 20% increase or MLD 2mm increase. |
| Fu VX | 2018 | 10.3171/2017.3.JNS162033 | Netherlands | 2002-2016 | Retrospective cohort study | 38 | To evalaute the clinical outcome, reproducibility of volumetric response patterns, and tumor control rate after administering a second GKRS to treat VS | T1 ceT1 T2 x | TV | Regression – TV ≥10% reduction Growth – TV >10% increase Stable – TV <10% change |
| Gawish A | 2023 | 10.1016/j.clon.2022.10.014 | Germany | 2007-2020 | Retrospective cohort study | 134 | To evaluate the long-term results of hypofractionated SRT applied in five fractions for VS | ceT1 x | Dimension Measures not clear | No threshold |
| Giordano M | 2020 | 10.1177/1971400919896253 | Germany | NR | Retrospective cohort study | 30 | To evalaute correlation between radiological tumor characteristics and the presence of edema, describe its MRI features and classify the different edema patterns | T1 ceT1 T2 x | TV MLD | Not analysed |

| | | | | | | | | | |
|---|---|---|---|---|---|---|---|---|---|
| Gonzalez-Darder JM | 2020 | 10.1016/j.wneu.2020.04.073 | Spain | 2007-2019 | Retrospective cohort study | 30 | To evalaute the prognostic value of MRI enhancement patterns to determine the risk of tumor regrowth after microsurgery | ceT1 1mm slice thickness | TV | Not analysed |
| Gonzalez-Orus Alvarez-Morujo RJ | 2014 | 10.1016/j.otorri.2014.01.002 | Spain | 1993-2013 | Retrospective cohort study | 73 | To determine the natural course of the VS, progression frequency, and mean growth, as well as determining the factors that could lead to potential growth | Contrast-enhanced y x | MLD | Growth – MLD ≥2mm, increase |
| Guo X | 2021 | 10.3389/fonc.2021.633350 | China | 2016-2019 | Retrospective cohort study | 452 | To identify risk factors associated with POH and reoperation following the resection of VS | Contrast-enhanced y x | MLD | Not analysed |
| Gupta A | 2020 | 10.4103/0028-3886.304069 | India | 2005-2015 | Retrospective cohort study | 294 | To document the outcomes and quality of follow-up compliance after planned subtotal, near-total and gross-total resection of VS | ceT1 x | TV MLD | Not analysed |
| Hasegawa T | 2021 | 10.1007/s11060-020-03622-5 | Japan | 1991-2013 | Retrospective cohort study | 203 | To evaluate the predictors of long-term tumor control following SRS for Koos grade 4 VS | ceT1 T2 x | TV MLD | Regression – TV ≥25% reduction Growth – TV ≥25% increase Stable – TV <25% reduction or increase; |
| Hentschel MA | 2021 | 10.1111/coa.13661 | Netherlands | 1990-2016 | Retrospective cohort study | 1217 | To develop a prediction model to predict VS growth for patients in a wait and scan strategy | ceT1 (preferred) or T2 x | MLD | Growth – MLD ≥25% increase |
| Higuchi Y | 2021 | 10.1016/j.wneu.2020.12.175 | Japan | 2010-2020 | Retrospective cohort study | 65 | To compare the growth potential of small remnants (< 1 cm3 ) after VS surgery with that of treatment-naïve small VS. | ceT1 1mm slice thickness | TV | Growth – TV ≥25% increase |
| Ho HH | 2018 | 10.1371/journal.pone.0 | Taiwan | 2011-2015 | Retrospective cohort study | 100 | To estimate the volume of VS by an ice cream cone formula using thin-sliced MRI | ceT1 hrT2$^{FIESTA}$ 0.8-2 mm slice thickness | TV | Not analysed |

| | | | | | | | | | | |
|---|---|---|---|---|---|---|---|---|---|---|
| | | 192411 | | | | | | | | |
| Hougaard D | 2014 | 10.1016/j.amjoto.2013.08.002 | Denmark | 2002-2010 | Retrospective cohort study | 72 | To determine the inter- and intraobserver variability in measuring VS size | T1<br>ceT1<br>0.5-0.7mm slice<br>T2 | TV | Not analysed |
| Hunter JB | 2016 | 10.1097/MAO.00000000000001219 | USA | 1995-2015 | Case series | 1296 | To characterize the risk and predictors of growth during observation of VS | ceT1<br>x | MLD | Growth – MLD ≥2mm |
| Hwang I | 2022 | 10.1007/s00330-021-08517-1 | South Korea | 2017-2019 | Retrospective cohort study | 35 | To evalaute the predictive role of pretreatment DCE-MRI parameters regarding the tumor response after GKRS in sporadic VS | ceT1<br>4-5mm slice thickness | TV | Reduction – TV >20% reduction |
| Itoyama T | 2022 | 10.1016/j.wneu.2022.07.058 | Japan | 2005-2019 | Retrospective cohort study | 67 | To evalaute the features associated with rapid growth of VS using radiomics analysis on MRI together with clinical factors | ceT1<br>x | MLD | No threshold |
| Jones A | 2024 | 10.1097/MAO.00000000000004239 | Australia | 1992-2020 | Case series | 657 | To determine if hypointense cochlear MRI CISS signal correlates with hearing outcomes in conservatively managed VS patients | $hrT2^{CISS}$<br>x | MLD | Not analysed |
| Kim BS | 2016 | 10.1007/s10143-016-0728-5 | South Korea | 2001-2014 | Retrospective cohort study | 17 | To analyse long-term clinical and radiological data focusing on NF2-related VS patients | ceT1<br>FLAIR<br>x | TV | Growth – TV >20% increase<br>Regression – TV >20 reduction<br>Stable – TV <20% change |

| | | | | | | | | | | |
|---|---|---|---|---|---|---|---|---|---|---|
| Kim H | 2015 | 10.1016/j.radonc.2015.03.031 | USA | 1994-2001 | Retrospective cohort study | 19 | To compare GKRS and LINAC SRS for VS | ceT1<br>T2<br>1mm slice thickness | TV | Not analysed |
| Kim J | 2022 | 10.3389/fonc.2022.996186 | South Korea | 1991-2021 | Retrospective cohort study | 33 | To evalaute the outcomes of SRS in NF2 population | ceT1<br>3mm slice thickness | TV | Recurrence – TV >10% increase |
| Kim JS | 2021 | 10.1007/s00701-021-04870-8 | South Korea | 2002-2019 | Retrospective cohort study | 118 | To evalaute the the growth rate of newly diagnosed VS and the related predictive factors for tumor growth | ceT1<br>x | MLD | Growth – MLD ≥2mm/year increase |
| Koetsier KS | 2021 | 10.1097/MAO.0000000000003313 | USA | 2003-2018 | Retrospective cohort study | 221 | To assess the efficacy and toxicity of proton radiotherapy in VS | ceT1<br>hrT2$^{CISS}$<br>hrT2$^{c\text{-}FIESTA}$ | TV<br>MLD | Growth – TV >20% increase<br>Regression – TV >20% reduction<br>Stable – TV <20% change |
| Kollmann P | 2020 | 10.1038/s41598-020-68489-y | Germany | NR | Retrospective cohort study | 8 | To compare three volumetric image analysis tools | ceT1<br>1mm slice thickness | TV | Not analysed |
| Kontorinis G | 2015 | 10.1007/s00405-014-3317-7 | UK | Since 1991 | Retrospective cohort study | 56 | To evaluate changes in hearing over time in patients with NF2 treated conservatively | ceT1<br>1mm slice thickness<br>FLAIR | MLD | Growth – MLD ≥2mm increase |
| Kranzinger M | 2014 | 10.1007/s00066-014-0630-4 | Austria | 2001-2007 | Case series | 29 | To provide long term follow-up details regarding the dynamics of tumor volumes and hearing preservation after hypofractionated SRS | ceT1<br>2mm slice thickness | TV | Growth – MLD ≥2mm increase |

| | | | | | | | | | | |
|---|---|---|---|---|---|---|---|---|---|---|
| Kujawa A | 2024 | 10.3389/fncom.2024.1365727 | UK | 2006-2019 | Retrospective cohort study | 168 | To evaluate automatic segmentation of VS | ceT1 0.5-4mm slice thickness T2 <4 mm slice thickness | TV | Not analysed |
| Larjani S | 2014 | 10.1371/journal.pone.0110823 | Canada | 2005-2011 | Retrospective cohort study | 63 | To determine whether pre-treatment growth rate of VS predict response to SRS | hrT2[c-FIESTA] 1.5mm slice thickness | TV | Growth – TV >20% increase over 12 month Regression – TV >20% reduction |
| Lawson McLean A | 2016 | 10.1007/s00701-016-2927-9 | Germany | NR | Retrospective cohort study | 40 | To compare radiographic data of female and male patients with NF2 | ceT1 0.5-0.7mm slice thickness | TV | Growth – TV ≥20% |
| Lee SA | 2024 | 10.1016/j.clineuro.2024.108402 | South Korea | 2017-2020 | Retrospective cohort study | 25 | To investigate the association between signal intensity on FLAIR images and audiovestibular findings in patients with VS | ceT1 3mm slice thickness | MLD | Not analysed |
| Li P | 2021 | 10.1016/j.clineuro.2020.106365 | China | 2006-2019 | Retrospective cohort study | 60 | To evaluate factors associated with growth pattern inconsistency in NF2 | ceT1 5mm slice thickness | TV | The growth pattern was defined as decreased if the relative AGR was <5% |
| Lim KH | 2024 | 10.1007/s00405-023-08410-1 | South Korea | 2006-2022 | Retrospective cohort study | 124 | To evaluate the association between MRI features and hearing status using a new radiomics technique | ceT1 2-4 mm slice thickness | TV | Not analysed |
| Liu Y | 2024 | 10.1088/1361-6560/ad2ee4 | China | NR | Retrospective cohort study | 60 | To evaluate automatic segmentation of VS | T1 ceT1 T2 [x] | TV | No threshold |

| | | | | | | | | | |
|---|---|---|---|---|---|---|---|---|---|
| Lo AWS | 2019 | 10.12809/hkjr1916934 | Hong Kong | 1999-2015 | Retrospective cohort study | 18 | To evaluate the predictive factors of pseudoprogression and treatment related toxicities in VS treated with FSRT | Contrast-enhanced<br>y<br>x | MLD | Pseudoprogression – MLD ≥2mm with subsequent stabilisation or regression of tumour size |
| Mahboubi H | 2023 | 10.3390/brainsci13101490 | USA | 2017-2019 | Retrospective cohort study | 109 | To evaluate the correlation between intra-operative assessment of residual tumor and early and follow-up imaging. | ceT1<br>FLAIR<br>x | TV | Not analysed |
| Marinelli JP | 2022 | 10.1093/neuonc/noab303 | USA | 1998-2018 | Retrospective cohort study | 952 | To evaluate the natural history of sporadic VS volumetric tumor growth, including long-term growth patterns following initial detection of growth | ceT1<br>T2 if not available<br>x | TV | Growth – TV ≥20% increase |
| Miller ME | 2017 | 10.1002/lary.26525 | USA | 1993-2004 | Retrospective cohort study | 220 | To determine the optimal postoperative MRI schedule and length of follow-up for patients undergoing microsurgical excision of VS | ceT1<br>y<br>x | MLD | No threshold |
| Morris KA | 2016 | 10.1259/bjr.20160110 | UK | NR | Retrospective cohort study | 46 | To compare the sensitivity of linear and volumetric measurements on MRI in detecting VS progression in patients with NF2 on bevacizumab treatment as well as the extent to which this depends on the size of the tumour | ceT1<br><3mm slice thickness<br>T2<br><1mm slice thickness | TV<br>MLD | ReiNS criteria<br><br>Partial response – TV ≥20% reduction<br>Progression – TV ≥20% increase<br>Stable – TV <20% change<br><br>(i) a change of +2 mm (progression) or -2 mm (response) in any consistent solitary serial linear measure of the tumour (1D 2 mm)<br>(ii) a change of +3 mm (progression) or -3 mm (response) in any consistent solitary serial linear measure of the |

| | | | | | | | | | | |
|---|---|---|---|---|---|---|---|---|---|---|
| | | | | | | | | | | tumour (1D 3 mm)<br><br>(i) a change of >10% (2D 10%)<br>(ii) a change of >20% (2D 20%)<br>(iii) a change of 3 mm or more (2D 3 mm). |
| Neve OM | 2023 | 10.1002/ohn.470 | Netherlands | NR | Retrospective cohort study | 185 | To validate the automated 2D diameter measurements of VS on MRI | ceT1<br>T2 if not available<br>x | MLD | Not analysed |
| Picry A | 2016 | 10.1002/lary.25976 | France | 1999-2013 | Retrospective cohort study | 18 | To evaluate the long-term growth rate of VS in NF2 patients based on volumetric measurements | ceT1, 2mm slice thickness | TV<br>MLD | No threshold |
| Pizzini FB | 2020 | 10.1097/MAO.0000000000002436 | Italy | NR | Retrospective cohort study | 66 | To compare the diagnostic accuracy of high resolution T2-WI and gadolinium-enhanced T1-weighted image sequences in quantitative evaluation of VS | ceT1<br>T2<br>x | MLD | Not analysed |
| Reddy CE | 2014 | 10.1017/S0022215114001315 | UK | 1995-2009 | Retrospective cohort study | 45 | To evaluate the natural course of VS 15 to 31 mm in diameter | T1<br>ceT1<br>x | MLD | Change – MLD ≥2mm |
| Ren Y | 2021 | 10.1177/0194599820978246 | USA | 2008-2018 | Retrospective cohort study | 151 | To identify preoperative radiographic predictors of hearing preservation after retrosigmoid resection of VS | ceT1<br>x | MLD | Not analysed |
| Rues D | 2018 | 10.1016/j.wneu.2018.04.149 | Germany | 1991-2015 | Retrospective cohort study | 355 | To evaluate the clinical and radiologic outcome of patients with VS treated with LINAC or CyberKnife-based SRS with respect to tumor control, preservation of serviceable hearing, and toxicity | ceT1<br>x | MLD | Growth – MLD >3mm increase |

| | | | | | | | | | |
|---|---|---|---|---|---|---|---|---|---|
| Saleh E | 2022 | 10.1097/MAO.0000000000003562 | Italy | 1994-2019 | Retrospective cohort study | 339 | To evaluate intracanalicular VS that were managed by wait and scan and to analyze the possible predictors of tumor growth and hearing deterioration throughout the observation period | Contrast-enhanced [x] [y] | MLD | Slow growth – MLD ≥2mm and <4mm/yr increase<br>Fast growth - MLD ≥4mm/yr increase<br><br>No growth – MLD <2mm change |
| Schneider T | 2016 | 10.1007/s00330-015-3895-9 | USA | 2003-2013 | Retrospective cohort study | 162 | To determine clinical outcome of patients with VS after treatment with fractionated SRT and single-session SRS by using 3D quantitative response assessment on MRI | ceT1<br>1-3mm slice thickness | TV | No threshold |
| Selleck AM | 2021 | 10.1097/MAO.0000000000003055 | USA | NR | Retrospective cohort study | 152 | To evaluate the natural growth history of VS utilizing volumetric measurements in an observed patient population | hrT2[CISS] [x] | TV<br>MLD | TV ≥20%<br>MLD ≥20% |
| Sethi M | 2020 | 10.1097/MAO.0000000000002448 | UK | 2005-2014 | Retrospective cohort study | 341 | To evaluate the conditional probability of VS growth at particular time-points, given a patient has not grown thus far | ceT1<br>1mm slice thickness [x] | MLD | Growth – MLD ≥2mm increase |
| Slane BG | 2017 | 10.1016/j.prro.2016.10.016 | USA | 2004-2011 | Retrospective cohort study | 56 | To compare the radiographic and clinical outcomes of patients with vestibular schwannomas treated with either single fraction SRS, or five fractions of hypofractionated SRT, or 25-30 fractions of conventionally fractionated SRT | ceT1 [x] | MLD | MLD change (≥20%) based on RECIST criteria |

| | | | | | | | | | | |
|---|---|---|---|---|---|---|---|---|---|---|
| Speckter H | 2019 | 10.1016/j.wneu.2019.08.193 | Dominican Republic | NR | Retrospective cohort study | 23 | To evaluate the texture features of routine MRI to predict tumor volume reduction and transient versus permanent tumor progression of VS treated by GKRS | ceT1 x | TV | No threshold |
| Sverak P | 2019 | 10.1177/0194599818809085 | USA | 2009-2018 | Retrospective cohort study | 17 | To evaluate the effectiveness and toxicity of bevacizumab in NF2 | ceT1 3mm slice thickness | TV | Positive radiographic response – TV ≥20% reduction<br>Radiographic progression – TV ≥20% increase<br>Stable – TV <19% change |
| Tai A | 2024 | 10.1016/j.clineuro.2024.108114 | USA | 2014-2022 | Case series | 99 | To evaluate whether the degree of the tumor's compression on the middle cerebellar peduncle influences FNO and EOR in medium to large VS | ceT1 x | MLD | Not analysed |
| Tang S | 2014 | 10.1097/MAO.0000000000000459 | USA | 1991-2012 | Retrospective cohort study | 88 | To compare different methods of measuring tumor growth after resection of VS and to identify predictors of growth | ceT1 3mm slice thickness | MLD<br>Maximum axial area<br>TV | Growth – MLD >1.4mm, >7 $mm^2$ for 2D, and TV >133 $mm^3$ |
| Tang X | 2018 | 10.21037/tcr.2018.08.15 | China | 2011-2015 | Retrospective cohort study | 561 | To evaluate hearing outcomes between multisession and single session GKRS in patients with VS and determine prognostic factors associated with hearing preservation | Contrast-enhanced x y | TV | No threshold |
| Taniguchi M | 2016 | 10.1016/j.wneu.2016.07.012 | Japan | 2008-2015 | Retrospective cohort study | 77 | To evaluate tumor-specific factors related to the association of hydrocephalus with small-to medium-sized VS | ceT1 0.8mm slice thickness<br><br>T2 3mm slice thickness | MLD | Not analysed |

| | | | | | | | | | |
|---|---|---|---|---|---|---|---|---|---|
| Tatagiba M | 2023 | 10.1093/noajnl/vdad146 | Germany | 2005-2011 | Retrospective cohort study | 901 | To compare the nuances in the treatment of VS by surgery and SRS | Contrast-enhanced<br>[x]<br>[y] | TV | Not analysed |
| Teggi R | 2014 | PMCID: PMC4025177 PMID: 24843223 | Italy | 2008-2011 | Retrospective cohort study | 64 | To evaluate vestibular function in a selected group of VS patients | ceT1<br>2mm slice thickness<br><br>$hrT2^{CISS}$<br>2mm slice thickness | TV<br>MLD | Not analysed |
| Teh SR | 2017 | 10.1017/S002221511600935X | Australia | NR | Retrospective cohort study | 12 | To determine intra- and inter-observer measurement variability of CPA tumours in a specialised institution | SPACE<br>[x] | MLD | Not analysed |
| Teixeira BCDA | 2021 | 10.1177/1971400920980165 | Brazil | 2015-2019 | Retrospective cohort study | 54 | To evaluate imaging characteristics of the tumour and inner-ear structures and to vestibulocochlear functional tests | $hrT2^{c-FIESTA}$<br>T2<br>FLAIR<br>[x] | MLD | Not analysed |
| Tikka T | 2018 | 10.1097/MAO.0000000000001962 | UK | 2000-2015 | Retrospective cohort study | 540 | To explore the nature of spontaneously regressing VS and identify possible predictive factors | ceT1, $hrT2^{c-FIESTA}$<br>[x] | MLD | Regression – MLD ≥2mm derease per annum<br><br>Rapid regression – MLD ≥4mm reduction per annum |
| Tomita Y | 2015 | 10.1016/j.wneu.2015.02.005 | Japan | 1998-2010 | Retrospective cohort study | 76 | To evaluate the radiographical tumor growth to elucidate factors possibly predicting growth or regrowth of their tumors. | ceT1<br>1-3mm slice thickness | TV | Significant TV change as >10% |
| Trau G | 2021 | 10.1007/s00405-020- | France | 2013-2018 | Retrospective cohort study | 35 | To evaluate the epidemiological, clinical, and radiological characteristics of regressive VS, based on | Post-contrast 2D spin-echo T1<br>[x] | TV | No threshold |

| | | | | | | | | | | |
|---|---|---|---|---|---|---|---|---|---|---|
| | | 06530-6 | | | | | volumetric measurements on MRI to define which regressions are significant and to look for a correlation between a shrinkage of the tumor and the medical history, and the presence of clinical symptoms | | | |
| Troude L | 2018 | 10.1016/j.wneu.2018.07.093 | France | 2003-2015 | Retrospective cohort study | 169 | To evaluate the long-term clinical and radiologic outcomes of patients harboring large VS treated with a facial nerve-sparing technique | ceT1 [x] | TV | Regrowth – TV >20% increase |
| Truong LUF | 2023 | 10.1007/s00405-022-07651-w | France | 2001-2019 | Retrospective cohort study | 78 | To determine whether the analysis of textural heterogeneity of VS on MRI at diagnosis was predictive of their radiological evolutivity | ceT1 [x] | TV<br>MLD | Progression – MLD >2mm or TV ≥20% increase |
| Turek G | 2023 | 10.1177/00034894231169341 | Poland | 2011-2021 | Retrospective cohort study | 64 | To identify possible predictors of facial nerve function and hearing preservation | ceT1 [x] | TV | Ineffective control – TV >10% increase following a 3-year follow-up |
| Wage J | 2021 | 10.1016/j.adro.2021.100687 | USA | 2004-2015 | Retrospective cohort study | 112 | To evaluate the long-term outcomes of patients with VS treated with GKRS with modern techniques, with attention to posttreatment tumor growth dynamics, dosimetric predictors, and late toxicities | Contrast-enhanced [x] [y] | MLD | Pseudoprogression – TV ≥10% increase occurring between 2 and 18 months |
| Wagner F | 2018 | 10.1007/s00066-018-1361-8 | Switzerland | 2010-2016 | Retrospective cohort study | 47 | To compare 3D-CISS sequences before and after primary SRS of unilateral VS to evaluate the effect of radiosurgery on the 3D-CISS signal intensities of cochlea and sacculus/utriculus | Post-contrast 3D T1W MPR [x] | TV | Not analysed |

| | | | | | | | | | | |
|---|---|---|---|---|---|---|---|---|---|---|
| Watanabe S | 2019 | 10.1007/s00701-019-03951-z | Japan | 1990-2015 | Retrospective cohort study | 402 | To evaluate the 5 years of follow-up of VS after SRS | ceT1<br>1-2mm slice thickness | TV<br>MLD | Growth – TV ≥125% or MLD ≥110% relative to the pre-treatment baseline<br><br>Regression – TV ≤75% and/or <MLD ≤90% |
| Wong RX | 2018 | 10.11622/smedj.2018107 | Singapore | 2007-2014 | Retrospective cohort study | 77 | To evaluate patients with VS who were treated using a linear accelerator-based SRS | ceT1<br>1mm slice thickness | MLD | RECIST criteria 1.1.<br><br>Progression – MLD 20% increase |
| Wu CH | 2019 | 10.1016/j.jns.2019.02.008 | Taiwan | 2007-2014 | Retrospective cohort study | 70 | To evaluate outcomes of VS patients who had acute sensorineural hearing loss early after radiosurgery | y<br>4mm slice thickness | TV<br>MLD | Not analysed |
| Yamada H | 2022 | 10.1002/lary.29834 | Japan | 2010-2016 | Retrospective cohort study | 31 | To determine the relationship between signal intensity on gadolinium-enhanced MRI and growth of VS | ceT1<br>x | TV | Growth – TV >100mm$^3$ /year |
| Yang HC | 2021 | 10.1016/j.radonc.2020.10.041 | Taiwan | 2006–2014 | Retrospective cohort study | 336 | To determine whether the radiomics analysis based on preradiosurgical MRI data could predict the pseudoprogression and long-term outcome of VS after GKRS | T1<br>ceT1<br>2.9–3.1mm slice thickness | TV | Regression – TV >10% reduction<br>Non-response – TV <10% change |
| Yeole U | 2022 | 10.1055/s-0041-1729977 | India | 2006-2016 | Retrospective cohort study | 34 | To evaluate single center experience of VS patients treated with SRS | ceT1<br>x | TV | Regression – TV >10% reduction<br>Stable – TV <10% change<br>Progression or failure of GKRS – TV >10% increase |
| Zhang Z | 2023 | 10.3389/fnins.2023.1207149 | China | 2019-2022 | Retrospective cohort study | 300 | To evaluate automatic segmentation of VS | ceT1<br>5mm slice thickness | TV | Not analysed |

ceT1 = contrast enhanced T1. CISS = constructive interference in steady state. CPA = cerebellopontine angle. EC = extracanalicular. FIESTA = fast imaging employing steady-state acquisition. GKRS = Gamma Knife radiosurgery. hrT2 = high resolution T2. IAC = internal auditory canal. IC = Intracanalicular. MLD = maximal linear diameter. NR = Not reported. TV = tumor volume. VS = vestibular schwannomas.

[x] = Slice thickness not specified

y = Sequence not specified
